# Intrathalamic morphometry in infants with congenital heart disease and infants born preterm

**DOI:** 10.64898/2026.03.16.26348485

**Authors:** Benjamin Clayden, Barat Gal-Er, Mirthe E.M. van der Meijden, Daniel Cromb, Siân Wilson, Kuberan Pushparajah, John Simpson, Christopher Kelly, Andrew T.M. Chew, Joseph V. Hajnal, Mary A. Rutherford, Jonathan O’Muircheartaigh, Chiara Nosarti, A. David Edwards, Serena J. Counsell, Alexandra F. Bonthrone

**Author notes:** Corresponding Author: Dr Alexandra Bonthrone, Research Department of Early Life Imaging, Centre for the Developing Brain, School of Biomedical Engineering & Imaging Sciences, King’s College London.

## Abstract

**Objective:** To compare intrathalamic morphometry in infants born preterm, with congenital heart disease (CHD) and typical controls and investigate associations with neurodevelopmental outcomes.

**Methods:** 592 infants underwent T2-weighted brain MRI: 107 CHD [gestational age at birth (GA) ≥37.00 weeks], 126 preterm (GA 23.00-36.86), 359 controls (GA ≥37.00). We used data-driven structural covariance analysis to derive 8 components of coordinated expansion and contraction within the thalamus. Permutation testing was used to test associations between intrathalamic morphometry and group (control, CHD, preterm birth <32 weeks GA), GA in infants born preterm and controls, cerebral oxygen delivery (CDO_2_) in infants with CHD, and neurodevelopmental outcomes at 18-24 months.

**Results:** Preterm infants born <32 weeks GA differed from infants with CHD and controls in 6 components encompassing most of the thalamus. Infants with CHD differed from controls in 2 components containing medial, ventricle-bordering and some anterior and ventrolateral thalamic areas. GA was associated with 7 components covering most of the thalamus, excepting the left posterior thalamus. CDO_2_ was not associated with intrathalamic morphometry. Right posterior thalamus morphometry was associated with motor scores in preterm infants born <32 weeks, but not in controls or infants with CHD.

**Interpretation:** Preterm infants born <32.00 weeks showed widespread morphometric changes across the thalamus, with alterations in the right posterior thalamus associated with motor outcomes at 18 months. Thalamic alterations in CHD were less widespread, confined to medial, ventrolateral, and ventricle-bordering tissues, which were not related to CDO_2_. Together, these findings suggest distinct thalamic phenotypes in prematurity and CHD.

## 1. Introduction

Infants with congenital heart disease (CHD) and those born preterm are at elevated risk of childhood neurodevelopmental impairments across motor, cognitive, and behavioral domains^1–7^. Both populations show brain alterations in infancy, including reduced volumes and altered development of cortical and subcortical regions^8–12^, changes in functional^13–16^ and structural connectivity^17–21^, and frequent white matter injury^22–24^. Alterations in the infant brain are associated with subsequent neurodevelopmental outcomes in both CHD^25,26^ and preterm^15,27–30^ cohorts. However, the mechanisms underlying altered brain development and impaired neurodevelopmental outcomes likely differ between these clinical populations. Infants with CHD experience reduced cerebral oxygen delivery (CDO_2_) as well as placental changes, the degree of which are associated with differences in brain development^9,26,31–34^. Infants born preterm experience multiple exposures including inflammation, gut microbiotal changes, stress, poor nutrition, and respiratory disease which are associated with brain changes^35–42^. Altered neonatal brain development and subsequent influences on neurodevelopmental outcomes in individuals with CHD and those born preterm have yet to be fully characterised.

Altered thalamic structure is reported in both preterm infants^8,11,28,43–46^ and infants with CHD^9,10,47,48^. Thalamus volumes are reduced in both populations^8–11,26,28,44,48,49^, and have been linked with deficits in motor^15,28,50^ and cognitive^15,25,26,28,29,50^ domains in childhood. In preterm infants, alterations in thalamic surface and shape are also reported^43,45,46^, although this has not been investigated in infants with CHD. However, there is limited literature directly comparing brain development in preterm infants and infants with CHD. Recent studies have shown differences in cortical scaling between these populations^51^, as well as distinct alterations in global connectivity metrics^52,53^. Another study has shown that regional structural morphometry across several structures including the whole thalamus differ between infants born <32 weeks GA and infants with CHD^27^. Despite alterations across the whole thalamus, it is unknown whether certain areas of the thalamus are more vulnerable to the effects of prematurity or CHD.

The thalamus is composed of around 40-60 nuclei with distinct functional, cytoarchitectural, and connectivity properties^54–57^. However, most previous studies on infants born preterm or with CHD have assessed the thalamus as a single region. Some evidence from deformation-based morphometry studies suggest that the thalamus may not be uniformly altered in these populations. Specific volumetric reductions to medial and anterior regions of the thalamus have been reported in infants with CHD^9^ and to the mediodorsal thalamus in preterm infants with diffuse white matter injury^58^. In preterm infants, an association between procedural pain and reduced volume of lateral portions of the thalamus has also been reported^41^. A recent study using a diffusion MRI-based parcellation of the thalamus reported no changes in relative volume of thalamic clusters in preterm birth, but larger relative medial cluster volumes in CHD^59^. To our knowledge, no study has assessed intrathalamic morphometry directly in neonates with CHD, infants born preterm, and controls.

Structural covariance (SC) is a neuroimaging analysis approach that identifies covariation in structural brain features across individuals^60,61^. When combined with data-driven methods such as independent component analysis (ICA)^62^, SC decomposes structural brain data into spatial components of coordinated expansion and contraction. In and neonatal research, these components are thought to reflect areas of coordinated maturation^63^. Previous studies have utilized SC analysis to investigate regional development across the brain in neonatal populations^64,65^, and altered brain structure in fetal and neonatal clinical populations^27,43,64–69^. SC has also been used in adult populations to investigate structural morphometry within specific brain regions, such as the hippocampus, striatum and insula^70–72^.

The aims of this study were to (i) use a data-driven SC approach to derive components of coordinated development within the thalamus in neonates; (ii) compare components between infants with CHD, infants born preterm, and typically developing infants ;(iii) investigate associations between components and GA at birth in infants born preterm and typical neonates ;(iv) assess associations between components and CDO_2_ in infants with CHD; and (v) characterise associations between components and neurodevelopmental outcomes at 18-22 months.

## 2. Methods

### 2.1 Ethical approval and informed consent

NHS Ethical approval was obtained (CHD: 07/H0707/105, 21/WA/0075; preterm and controls: 14/LO/1169). Written informed parental consent was obtained before MRI scan and neurodevelopmental assessments.

### 2.2 Recruitment

Recruitment was previously reported in Wilson and colleagues^27^. Data from control and preterm infants were from the developing Human Connectome Project (dHCP) 3^rd^ neonatal data release^73^ (https://nda.nih.gov/edit_collection.html?id=3955), a sample of infants born 23.00-44.00 weeks GA from Greater London.

The typically developing control sample included 359 infants. Exclusion criteria were GA <37.00 weeks and a cognitive and motor composite score below 70 (>2 SD below the mean on the Bayley Scales of Infant and Toddler Development, Third Edition^74^ (Bayley-III)) at 18 months. Infants were also excluded if they had prenatal maternal exposure to COVID19, or had a first-degree relative with a psychiatric condition, autism or, ADHD.

The preterm sample consisted of 126 infants born 23.00-36.86 weeks GA. We further categorised infants born preterm into early preterm birth (GA <32.00, n=60) and late preterm birth (GA 32.00-36.86, n=66).

The CHD sample included 107 infants with critical or serious CHD, defined as requiring cardiac catheterisation or surgery before 1-year postnatal age^27,75^, recruited as fetuses or neonates from St Thomas’ Hospital, London. Exclusion criteria were a suspected/confirmed chromosomal abnormality/congenital syndrome, previous neonatal surgery, or birth <37.00 weeks GA.

Exclusion criteria for all infants included major lesions on MRI (e.g. arterial ischemic stroke, parenchymal hemorrage, >10 foci of punctate white matter injury), poor MR image quality due to motion, or incomplete/no T2-weighted imaging.

### 2.3 MR data acquisition and reconstruction

All infants underwent brain MRI between 37.00-45.14 weeks post-menstrual age (PMA) on a Phillips 3T Achieva system (Best, Netherlands) situated on the neonatal unit at St. Thomas Hospital, London. Infants were scanned using a dedicated neonatal brain imaging system^76^ with a custom 32-channel receive neonatal head coil (Rapid Biomedical GmbH, Rimpar, DE). Hearing protection was applied with ear putty and earmuffs (President Putty, Coltene Whaledent, Mahwah, NJ, USA), and inflatable pads (Pearltec, Zurich CH) were used for positioning and to limit head motion. Infants were imaged during natural sleep. Before scanning, infants were fed, swaddled, and positioned in a vacuum jacket. Scans were supervised by a neonatal nurse and/or pediatrician experienced in neonatal MR procedures, with monitoring of heart rate, oxygen saturation, and body temperature throughout acquisition.

T2-weighted Turbo Spin Echo MRI were acquired in the axial and sagittal planes using the dHCP neonatal acquisition protocol^73,76^: Slice thickness=1.6mm, in-plane resolution=0.89mm, TR=12s, TE=156ms, flip angle=90°, FOV=145×145×108mm^3^, SENSE factor 2.11 (axial) and 2.58 (sagittal). Specialised neonatal pipelines for motion-correction^77^ and super-resolution reconstruction^78^ were used to reconstruct volumes using data from both acquisitions (reconstructed voxel size= 0.5mm^3^ isotropic).

Infants with CHD underwent velocity sensitized phase contrast angiography to measure cerebral blood flow (single-slice T1-weighted fast field echo sequence, resolution= 0.6×0.6×4.0mm, TR=6.4ms, TE=4.3ms, flip angle=10°, FOV=100×100mm^2^, repetitions=20, maximal encoding velocity=140cm/s). Images were acquired in a plane perpendicular to both internal carotids and basilar arteries at the level of the sphenoid bone^79^. Good quality phase contrast angiography was available in 96 (90%) infants with CHD.

### 2.4 Neurodevelopmental assessment

Infants were invited to attend a neurodevelopmental follow-up assessment as close to 18 months (dHCP) or 22 months (CHD) corrected age as possible (range=17-40 months). Bayley III^74^ were administered by a developmental pediatrician or psychologist to obtain cognitive, language, and motor composite scores [test mean (SD) =100 (15)]. Cognitive, language and motor composite scores were available for 357 controls, 72 infants with CHD and 47 early preterm infants. Additionally, three infants with CHD had cognitive and motor composite scores available, and one infant with CHD had cognitive composite scores alone. Parents were also asked to complete a questionnaire asking whether English was spoken as an additional language by either parent (data available in control=342, CHD=72, early preterm=45).

### 2.5 Cerebral oxygen delivery

Cerebral blood flow (CBF) was calculated from phase contrast angiography using previously published methods^26,33,34^. Briefly, the right/left internal carotid and basilar arteries were manually segmented using Segment v2.0 R4800^80^. Flow was then extracted and summed across the vessels. Haemoglobin (Hb) measurements were extracted from clinical notes at a median of 2 days (IQR=1-4) before scan. Preductal oxygen saturation (SaO_2_) was measured at time of scan with a Masimo Radical-7 monitor (Masimo Corp, Irvine, CA) applied to the right hand.

Cerebral oxygen delivery (CDO_2_) was calculated using the following formula^81^:

CDO_2_ (mLO_2_/min) = SaO_2_ x Hb (g/dL) x 1.36 x CBF (mL/min)

Where 1.36 is the amount of oxygen bound per gram of haemoglobin at one atmosphere pressure (Hüfner’s constant)^31^.

### 2.6 Additional data collected

Birthweight z-scores were calculated for all infants using the UK-WHO 1990 normative data from 23 weeks GA^82^.

Index of Multiple Deprivation (IMD) was calculated using the 2019 data release for postcode at birth and reported as rank and quintiles (most to least deprived). IMD is a composite measure of socioeconomic status in England encompassing factors including income, employment, education, health, and crime. IMD was missing for 5 infants (control=2, CHD=3).

### 2.7 MRI processing

Processing is previously described by Wilson and colleagues^27^. Images were processed using the dHCP structural pipeline^83^ which includes bias correction^84^, skull-stripping^85^, and segmentation into tissue classes using a neonatal-specific expectation maximisation algorithm^83,86^. Skull-stripped, bias-field corrected images underwent multistep non-linear registration to a template from the dHCP extended atlas corresponding to the infant’s PMA at scan^87^ (https://gin.g-node.org/BioMedIA/dhcp-volumetric-atlas-groupwise), and then to a central template corresponding to 40 weeks GA using the symmetric diffeomorphic registration (SyN) algorithm with a cross-correlation metric from Advanced Normalization Tools 3.0 (ANTs)^88^. Log-Jacobian determinant maps were generated for each infant from the registration warps from individual to the 40-week GA atlas space and resampled to 1mm^3^. Log-Jacobian values represent expansion and contraction of brain regions during registration^89^.

### 2.8 Independent Component Analysis (ICA) of Jacobian determinants

We applied a canonical ICA algorithm^90^ (canICA) to the log-Jacobian determinants, implemented in Python with the nilearn package^91^. The algorithm breaks down input data into separate, maximally independent components with non-Gaussian distributions. CanICA was applied with a thalamic mask to identify independent components (ICs) of coordinated expansion and contraction within the thalamus. The optimal number of ICs (n=8) was chosen based on the variance explained by components (Supplementary Table 1, Supplementary Figure 1), and visual inspection of the spatial distribution of components.

FMRIB Software Library’s (FSL) General Linear Model^92,93^ (GLM) was used to extract a weight for each IC in each infant, which captures how strongly an IC maps onto an individual’s thalamic morphometry.

### 2.9 Total thalamus and lateral ventricle volumes

Total thalamus and lateral ventricle volumes were calculated from segmentations produced by the dHCP structural pipeline^83^.

### 2.10 Statistical comparisons

All statistical comparisons were carried out in Python (v3.13).

Multiple linear regressions with permutation testing (n=10,000) were used for all analyses. We investigated differences in IC weightings between typically developing control infants, infants with CHD and infants in the early preterm group, covarying for PMA at scan, birthweight z-score, infant sex and whether the infant was part of a multiple birth. For all comparisons between infants with CHD and typically developing controls, postnatal age (PNA) at scan was also included as a covariate.

To investigate if differences in IC weightings between groups were driven by total thalamus or ventricular volumes, analyses were run additionally covarying for total thalamus or lateral ventricle volume.

In order to characterise the relationship between degree of prematurity and changes in thalamic morphometry, we investigated associations between GA at birth and IC weightings across control, late preterm, and early preterm infants (n=485), adjusting for PMA at scan, birthweight z-score, infant sex, and multiple birth status.

We also assessed if CDO_2_ was associated with IC weightings in infants with CHD adjusting for PMA at scan, PNA at scan, birthweight z-score, infant sex, and multiple birth status.

To characterise the relationship between IC weightings and neurodevelopmental outcomes, we assessed whether the interaction between IC weights and infant group (control, CHD, early preterm) was associated with cognitive, language, or motor composite scores adjusting for IMD, PMA at scan, birthweight z-score, infant sex, and multiple birth status. Language analyses were additionally adjusted for whether English was an additional language spoken by at least one parent.

We investigated differences in total thalamus and lateral ventricle volumes between groups, adjusting for PMA at scan, birthweight z-score, infant sex, and multiple birth status. For comparisons between infants with CHD and typically developing controls, PNA at scan was also included as a covariate.

*p*-values were corrected for multiple comparisons using Holm-Bonferroni correction and are reported as *p*_FWE_.

When a significant interaction was identified in neurodevelopmental outcome analyses, post-hoc analyses were run assessing the relationship between IC weightings and outcome scores separately in each group adjusting for PMA at scan, PNA at scan, birthweight z-score, infant sex, multiple birth status, and IMD. These analyses were repeated additionally covarying for total thalamus or lateral ventricle volume. Analyses were also run to characterise the relationship between total thalamus and lateral ventricle volumes with significant outcome scores in each group, covarying for PMA at scan, PNA at scan, birthweight z-score, infant sex, multiple birth status, and IMD.

## 3. Results

### 3.1 Demographic information

Demographic information is provided in Table 1.

**Table 1.** Demographics at base and follow-up.

| **Table 1. Demographics at base and follow-up** |  |  |  |  |
| --- | --- | --- | --- | --- |
|  | **Control**<br>**(n = 359)** | **CHD**<br>**(n = 107)** | **Late**<br>**Preterm**<br>**(n = 66)** | **Early**<br>**Preterm**<br>**(n = 60)** |
| **Neonatal Characteristics** |  |  |  |  |
| GA at Birth, weeks,<br>median [IQR] | 40.14 [39.14-41] | 38.43 [0.72] | 35.00 [2.43] | 28.71 [4.07] |
| PMA at Scan, weeks,<br>median [IQR] | 41.43 [40.14-42.86] | 39.14 [1.14] | 40.86 [2.65] | 41.36 [2.72] |
| Birthweight Z-Score,<br>weeks, median [IQR] | -0.19 [-0.79-0.46] | -0.34 [1.22] | -0.17 [1.55] | -0.41 [1.40] |
| Female, n (%) | 185 (51.53%) | 59 (55.14%) | 38 (57.58%) | 33 (55.00%) |
| Singleton, n (%) | 346 (96.38%) | 101 (94.39%) | 42 (63.64%) | 42 (70.00%) |
| CDO<sub>2</sub> (mLO<sub>2</sub>/min). |  | 1644 [1432- |  |  |
| Median [IQR] |  | 2094] |  |  |
| **Neurodevelopmental follow-up characteristics** |  |  |  |  |
| Corrected Age at Assessment, months, median [IQR] | 18.46 [18.07-19.18] | 22.32 [22.06-23.8] | 18.44 [18.3-19.56] | 18.53 [18.22-19.33] |
| Cognitive composite score, mean [SD] | 101.87 [10.69] | 93.99 [10.92] | 101.5 [12.67] | 96.06 [11.98] |
| Motor composite score, mean [SD] | 103.75 [8.66] | 96.99 [10.14] | 101.42 [9.08] | 96.3 [13.25] |
| Language composite score, mean [SD] | 99.69 ([4.97] | 94.06 [17.12] | 96.6 [18.05] | 96.25 [18.19] |
| Parent first language [English], n (%) | 180 [52.94%] | 48 [72.73%] | 28 [58.33%] | 34 [77.27%] |
| Index of multiple deprivation score, median [IQR] | 12030 [8158-18728] | 18982 [9538-24669] | 15964 [11756-21424] | 14649 [9373-21776] |
| Index of multiple deprivation (quintiles) | N (%) |  |  |  |
| First (most deprived) | 52 (14.7) | 11 (14.9) | 5 (10.0) | 5 (10.6) |
| Second | 145 (40.9) | 16 (21.6) | 12 (24.0) | 14 (29.8) |
| Third | 78 (22.0) | 13 (17.6) | 16 (32.0) | 14 (29.8) |
| Fourth | 52 (14.7) | 17 (23.0) | 13 (26.0) | 3 (6.4) |
| Fifth | 28 (7.9) | 17 (23.0) | 4 (8.0) | 11 (23.4) |
| Demographics of infant groups at follow up includes infants with at-least one composite score recorded. |  |  |  |  |

### 3.2 Intrathalamic structural covariance components

All ICs were either bilateral or had a complimentary contralateral component (Figure 1). Components covered almost the entirety of the thalamus, including posterior, anterior, ventral, dorsal, medial, and lateral areas (Table 2).

**Figure 1:**
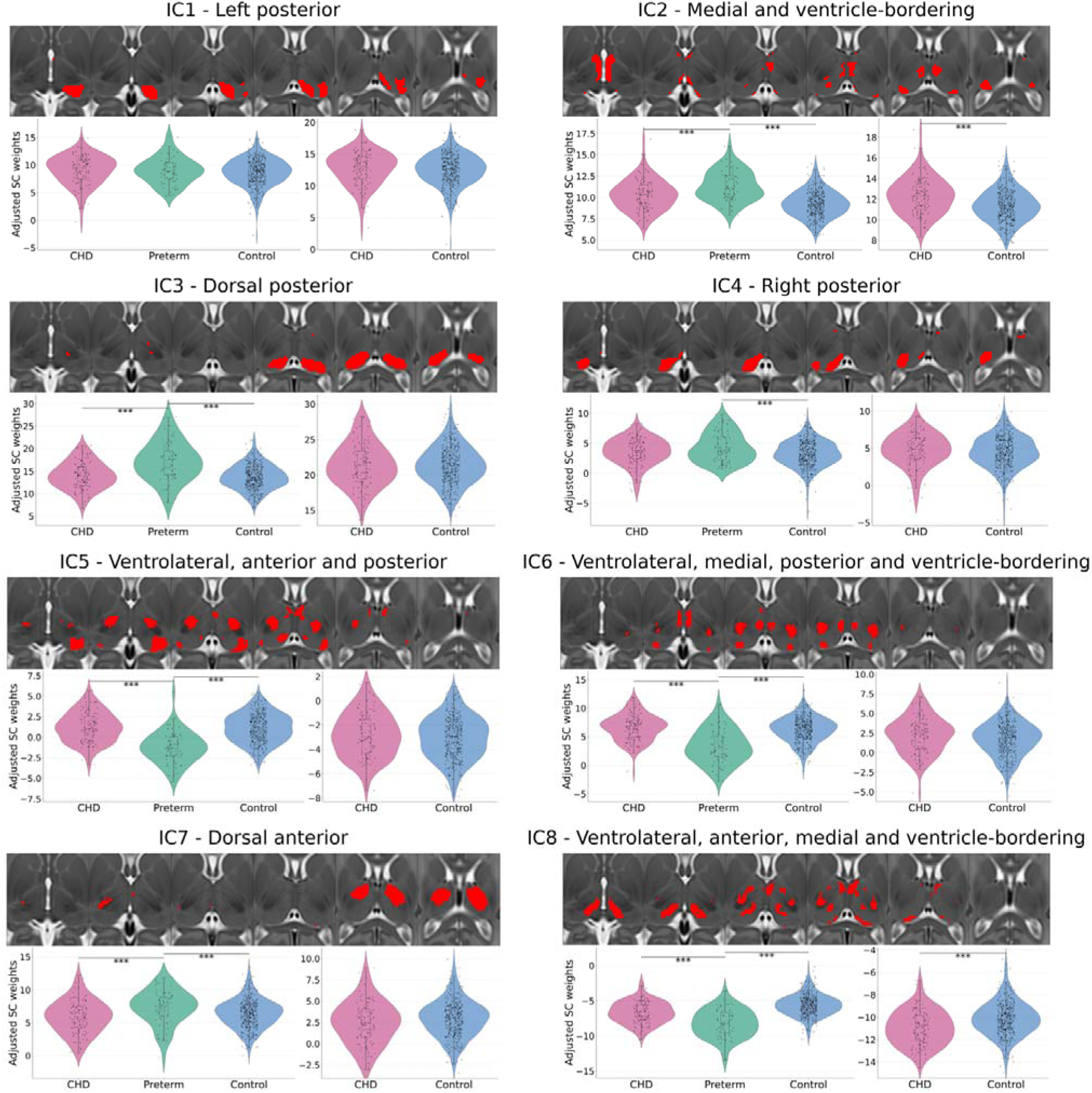
Intrathalamic components extracted using ICA. Each component is shown in the axial plane overlaid on the dHCP extended atlas 40-week template. IC weights adjusting for PMA at scan, birthweight, sex and whether the infant was part of a multiple birth are shown for each group. CHD v Control violin plots are additionally adjusted for PNA at scan. *** p_FWE_<0.001, ** p_FWE_<0.01, * p_FWE_<0.05

**Table 2.** Anatomical description of derived intrathalamic independent components (IC)

|  |  |
| --- | --- |
| IC1 | Left posterior |
| IC2 | Medial and ventricle-bordering |
| IC3 | Dorsal posterior |
| IC4 | Right posterior |
| IC5 | Ventrolateral, anterior bilaterally, and posterior (left>right) |
| IC6 | Ventrolateral, medial, posterior and ventricle-bordering |
| IC7 | Dorsal anterior |
| IC8 | Ventrolateral, anterior, medial and ventricle-bordering |

### 3.2. Intrathalamic morphometry differences between groups

Early preterm infants differed from both CHD and typically developing controls in six components (IC 2,3,5,6,7,8), encompassing dorsal, ventrolateral, medial, anterior, posterior, and ventricle-bordering thalamic areas (*p*_FWE_≤0.003, Table 3, Figure 1). Early preterm infants were also different from controls in IC4 (right posterior thalamus) (*p*_FWE_=0.003). CHD and controls differed in two components (IC2, IC8) (*p*_FWE_<0.001), containing medial and ventricular-bordering areas (IC8 also containing some anterior and ventrolateral regions). Weightings in IC1 (left posterior thalamus) did not differ between groups (*p*_FWE_≥0.249).

**Table 3.** Differences in intrathalamic independent component (IC) weightings between control infants, infants with CHD and preterm infants.

|  |  |  |  |  |  |  |  |  |  |
| --- | --- | --- | --- | --- | --- | --- | --- | --- | --- |
|  | CHD Vs Control |  |  | Early Preterm Vs Control |  |  | CHD Vs Early Preterm |  |  |
| Intrathalamic IC | b | *p* | *p*<sub>FWE</sub> | b | *p* | *p*<sub>FWE</sub> | b | *p* | *p*<sub>FWE</sub> |
| *IC weight ~ Group + Covariates* |  |  |  |  |  |  |  |  |  |
| 1 | -0.365 | 0.260 | 0.647 | -0.422 | 0.249 | 0.249 | 0.693 | 0.209 | 0.418 |
| 2 | -1.343 | <0.001 | **<0.001** | -2.377 | <0.001 | **<0.001** | 1.601 | <0.001 | **0.001** |
| 3 | -0.126 | 0.729 | 0.729 | -4.034 | <0.001 | **<0.001** | 3.651 | <0.001 | **<0.001** |
| 4 | -0.400 | 0.162 | 0.647 | -1.206 | 0.001 | **0.002** | 0.021 | 0.969 | 0.969 |
| 5 | -0.306 | 0.186 | 0.647 | 2.756 | <0.001 | **<0.001** | -2.990 | <0.001 | **<0.001** |
| 6 | -0.755 | 0.009 | 0.052 | 3.350 | <0.001 | **<0.001** | -3.399 | <0.001 | **<0.001** |
| 7 | 0.655 | 0.024 | 0.118 | -1.080 | 0.001 | **0.003** | 2.043 | <0.001 | **<0.001** |
| 8 | 1.151 | <0.001 | **<0.001** | 2.930 | <0.001 | **<0.001** | -1.448 | <0.001 | **<0.001** |

| *IC weight ~ Group + Thalamus Volume + Covariates* |  |  |  |  |  |  |  |  |  |
| --- | --- | --- | --- | --- | --- | --- | --- | --- | --- |
| 1 | -0.529 | 0.117 | 0.533 | -0.792 | 0.263 | 0.450 | -0.065 | 0.942 | 1.000 |
| 2 | -1.078 | <0.001 | **<0.001** | -1.343 | 0.001 | **0.006** | 0.005 | 0.991 | 1.000 |
| 3 | -0.532 | 0.152 | 0.533 | -4.611 | <0.001 | **<0.001** | 3.039 | 0.008 | 0.053 |
| 4 | -0.479 | 0.107 | 0.533 | -0.769 | 0.225 | 0.450 | 0.541 | 0.458 | 1.000 |
| 5 | -0.234 | 0.336 | 0.673 | 2.276 | <0.001 | **<0.001** | -2.147 | <0.001 | **0.002** |
| 6 | -0.904 | 0.003 | **0.017** | 2.314 | <0.001 | **0.001** | -1.945 | 0.012 | 0.074 |
| 7 | 0.074 | 0.806 | 0.806 | -2.563 | <0.001 | **<0.001** | 1.751 | 0.031 | 0.156 |
| 8 | 1.024 | <0.001 | **<0.001** | 1.326 | 0.002 | **0.007** | 0.141 | 0.786 | 1.000 |
| *IC weight ~ Group + Lateral Ventricle Volume + Covariates* |  |  |  |  |  |  |  |  |  |
| 1 | -0.539 | 0.078 | 0.282 | 0.522 | 0.139 | 0.139 | -0.677 | 0.211 | 0.422 |
| 2 | -1.215 | <0.001 | **<0.001** | 2.302 | <0.001 | **<0.001** | 1.580 | <0.001 | **<0.001** |
| 3 | -0.085 | 0.811 | 0.811 | 4.037 | <0.001 | **<0.001** | 3.649 | <0.001 | **<0.001** |
| 4 | -0.500 | 0.070 | 0.282 | 1.265 | <0.001 | **0.001** | 0.032 | 0.948 | 0.948 |
| 5 | -0.291 | 0.212 | 0.425 | -2.786 | <0.001 | **<0.000** | -3.005 | <0.001 | **<0.001** |
| 6 | -0.820 | 0.004 | **0.023** | -3.336 | <0.001 | **<0.001** | -3.411 | <0.001 | **<0.001** |
| 7 | 0.710 | 0.014 | 0.072 | 1.074 | 0.002 | **0.003** | 2.047 | <0.001 | **<0.001** |
| 8 | 1.154 | <0.001 | **<0.001** | -2.964 | <0.001 | **<.001** | -1.456 | <0.001 | **<0.001** |
| Covariates: for PMA at scan, birthweight, sex and whether the infant was part of a multiple birth. Comparisons between control and infants with CHD were also adjusted for PNA at scan. Significant associations are in bold. |  |  |  |  |  |  |  |  |  |

Adjusting for lateral ventricle volume did not modify the alterations identified in early preterm infants compared to infants with CHD or controls (Table 3). IC2 and IC8 remained significantly different between infants with CHD and controls and IC6 (ventrolateral, medial, ventricle-bordering, and posterior regions) was additionally different (*p*_FWE_≤0.023).

When adjusting for total thalamus volume, early preterm infants remained significantly different from controls in 6/8 components (*p*_FWE_≤0.007, Table 3). However, IC4 was no longer significantly different between groups. Only IC5 (containing ventrolateral, posterior and anterior areas) remained significantly different between early preterm infants and those with CHD (*p*_FWE_=0.002). IC2 and IC8 remained significantly different in infants with CHD compared to controls, and IC6 (ventrolateral, medial, ventricle-bordering and posterior regions) was additionally altered (*p*_FWE_≤0.017).

### 3.3 Association between GA at birth and intrathalamic morphometry

GA at birth was associated with seven ICs (*p*_FWE_≤0.008, Table 4, Figure 2). Weightings in IC1 (left posterior thalamus) were not associated with GA at birth (*p*_FWE_=0.106).

**Figure 2:**
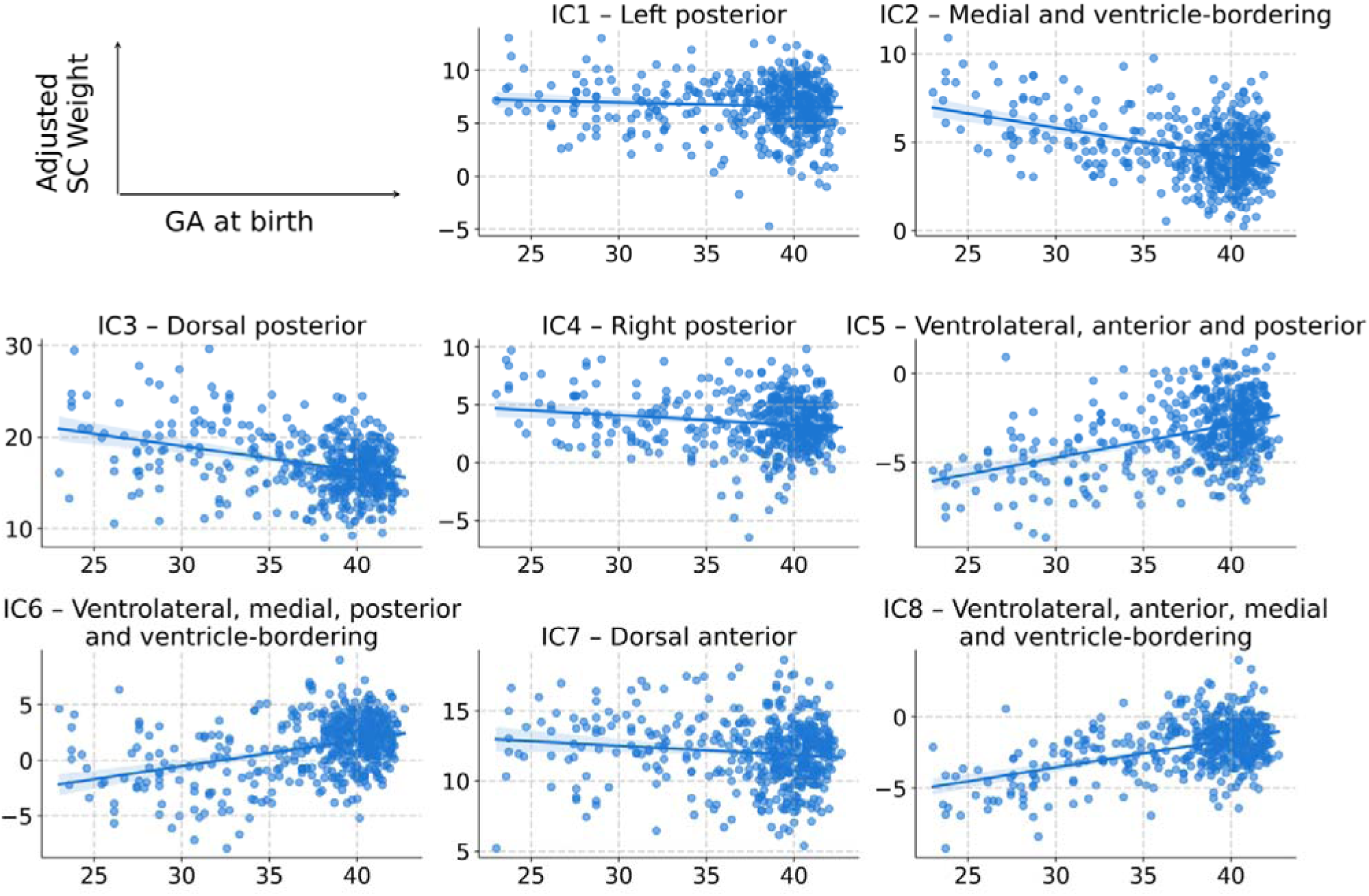
Association of GA at birth and weightings for each IC adjusting for PMA at scan, birthweight, sex and whether the infant was part of a multiple birth.

**Table 4.** Association between gestational age at birth and intrathalamic independent component (IC) weights.

|  |  |  |  |
| --- | --- | --- | --- |
|  | **b** | *p* | *p*<sub>FWE</sub> |
| IC1 | -0.046 | 0.106 | 0.106 |
| IC2 | -0.190 | <0.001 | **<0.001** |
| IC3 | -0.315 | <0.001 | **<0.001** |
| IC4 | -0.099 | <0.001 | **<0.001** |
| IC5 | 0.219 | <0.001 | **<0.001** |
| IC6 | 0.275 | <0.001 | **<0.001** |
| IC7 | -0.077 | 0.004 | **0.008** |
| IC8 | 0.231 | <0.001 | **<0.001** |
| adjusting for PMA at scan, birthweight, sex and whether the infant was part of a multiple birth. Significant associations are in bold. |  |  |  |

### 3.4 Association between CDO_2_ and intrathalamic morphometry

There were no significant associations between CDO_2_ and IC weightings (*p*_FWE_≥0.444) (Supplementary Table 2).

### 3.5 Relationship between intrathalamic morphometry and neurodevelopmental outcomes

The interaction between IC4 (right posterior thalamus) weightings and infant group (early preterm vs control) was associated with motor composite scores (*p*_FWE_=0.035, Figure 3, Table 5). Post-hoc analysis revealed motor composite scores were associated with IC4 weights in early preterm infants (*p*=0.037) but not controls (*p*=0.675) or infants with CHD (*p*=0.555) (Supplementary Table 3). These results were not altered when either total thalamus volume (early preterm; *p*=0.023; controls; *p*=0.638 CHD; *p*=0.581) or total lateral ventricle volume (early preterm; *p*=0.043; controls; *p*=0.331 CHD; *p*=0.608) were included as an additional covariate (Supplementary Table 3). No other group-IC weighting interactions were associated neurodevelopmental outcomes (Table 5).

**Figure 3:**
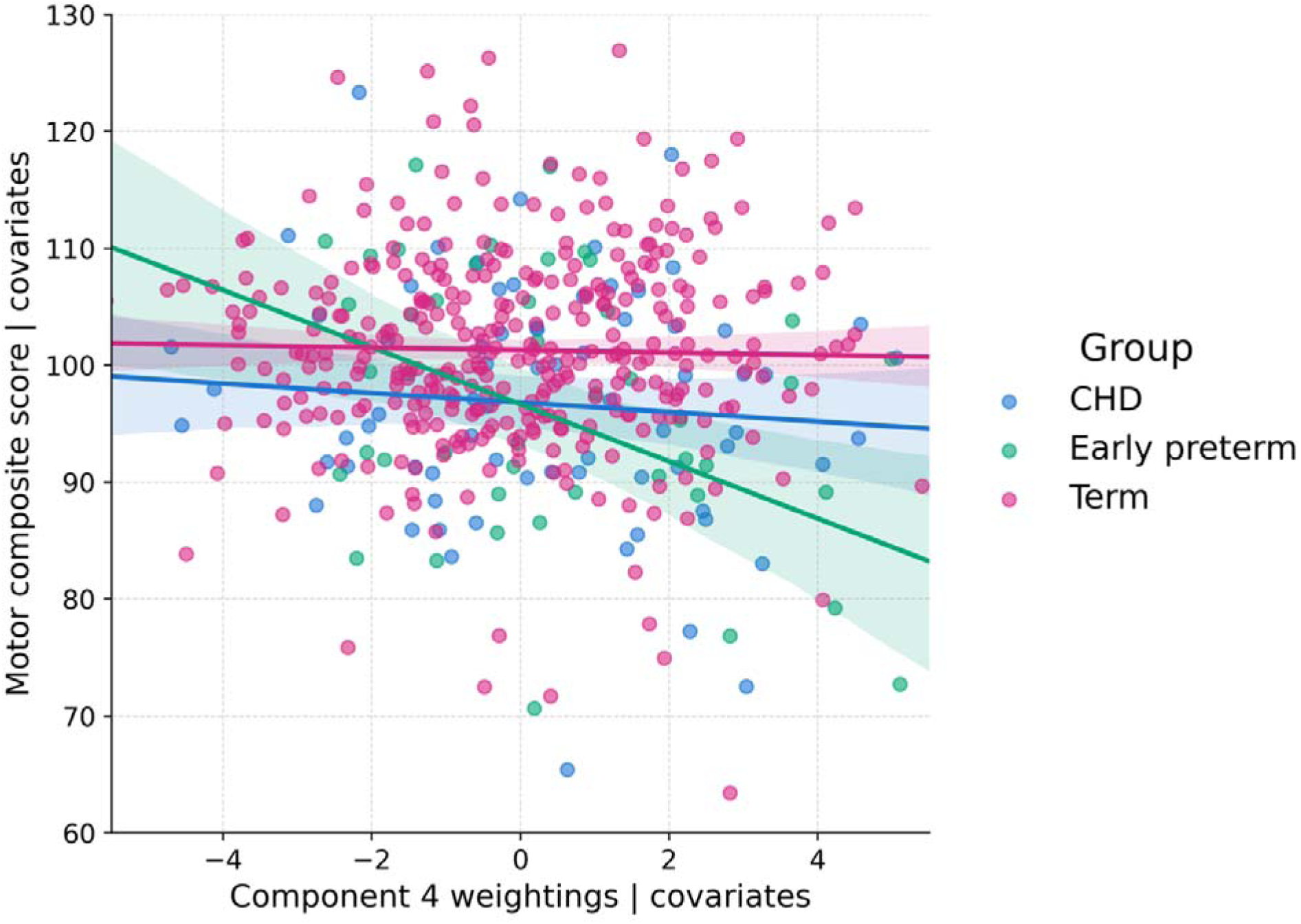
Association between weightings in IC4 and motor composite scores in early life adjusting for IMD, PMA at scan, birthweight, sex and whether the infant was part of a multiple birth.

**Table 5.**
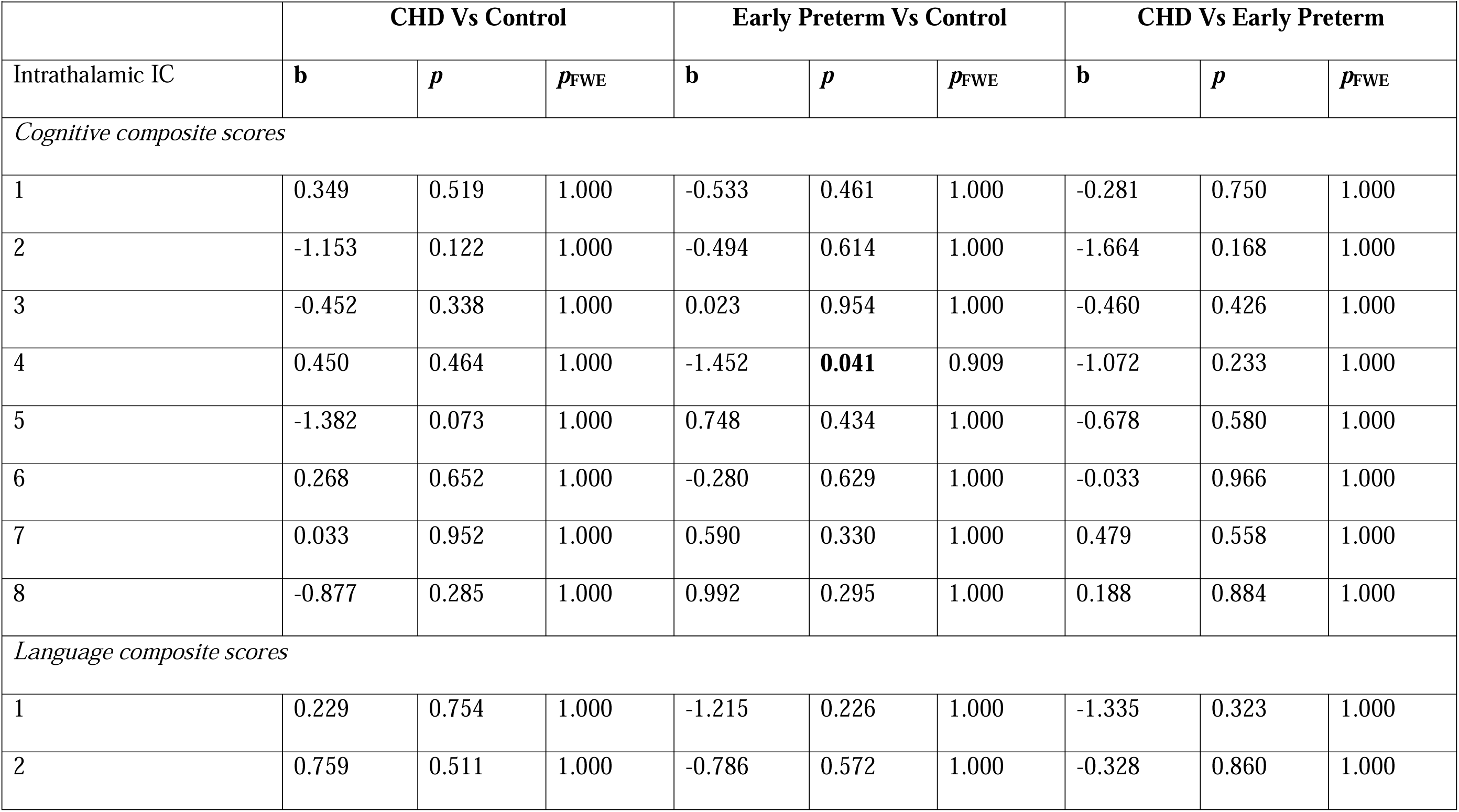
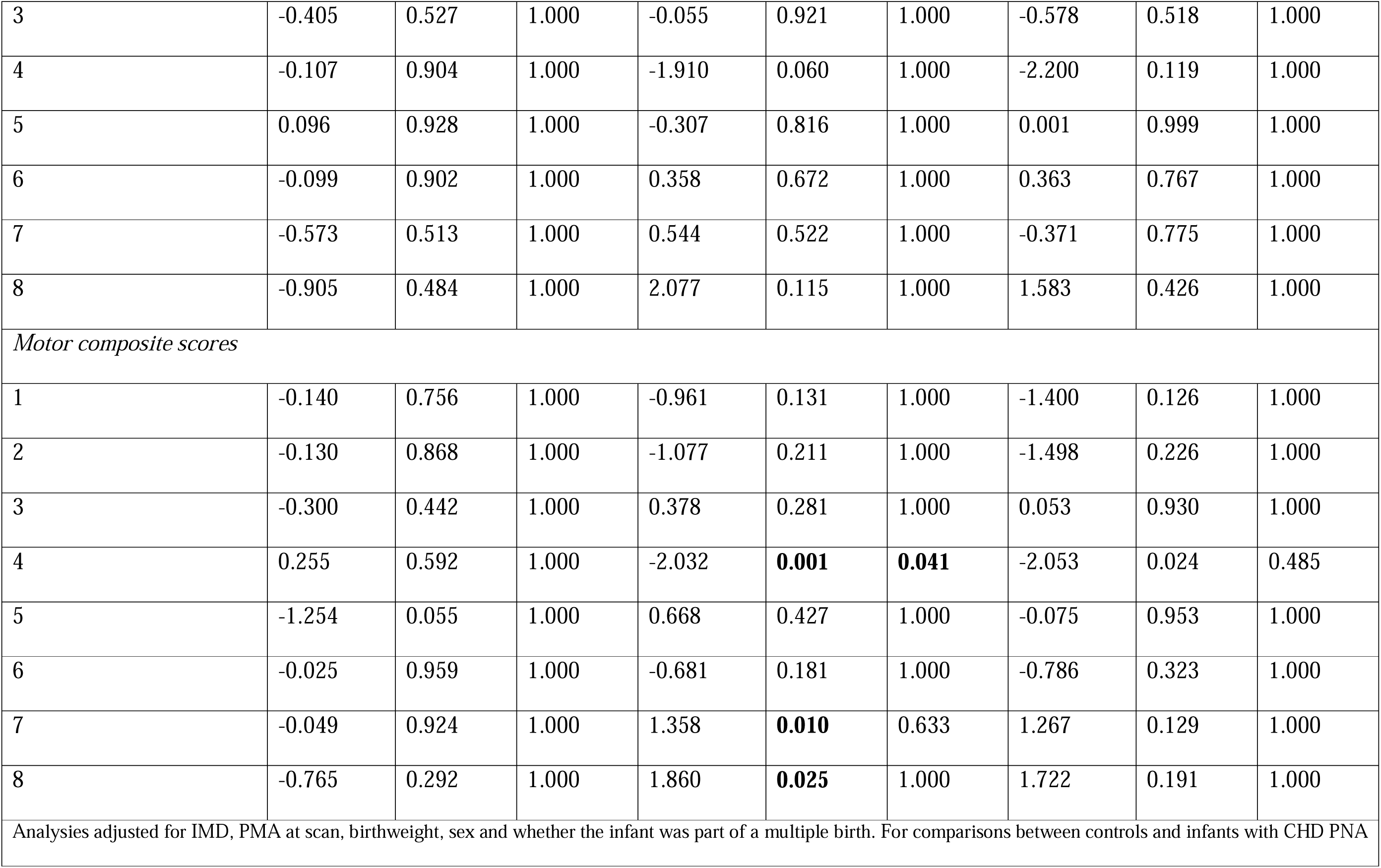
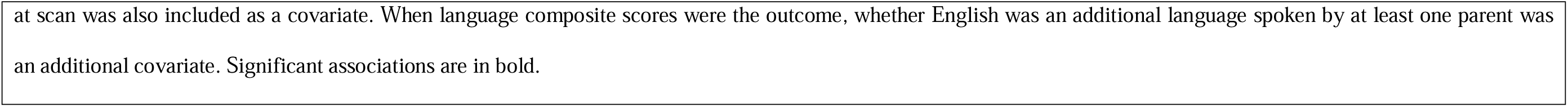
Interaction effect of infant group on the relationship between intrathalamic independent component (IC) weights and neurodevelopmental outcomes.

### 3.6 Total thalamus and lateral ventricle volumes

Total thalamus volume was significantly different between controls, infants with CHD and infants in the early preterm group (*p*<0.001) (Table 6). Total lateral ventricle volume did not differ between groups (*p*≥0.130) (Table 6).

**Table 6.** Total thalamus and lateral ventricle volumes in each infant group.

|  |  |  |  |  |  |  |  |  |
| --- | --- | --- | --- | --- | --- | --- | --- | --- |
| Control | CHD | Early Preterm | CHD vs Control |  | Early Preterm vs Control |  | CHD vs Early Preterm |  |
| Median [IQR] | Median [IQR] | Median [IQR] | t | $p_{\text{FWE}}$ | t | $p_{\text{FWE}}$ | t | $p_{\text{FWE}}$ |
| *Thalamus volume* |  |  |  |  |  |  |  |  |
| 9128 [8480-9864] | 7793 [7364-8261] | 5007 [3992-5597] | 5.972 | **<0.001** | 33.5 | **<0.001** | 13.8 | **<0.001** |
| *Lateral ventricle volume* |  |  |  |  |  |  |  |  |
| 4620 [3806-5570] | 4446 [3511-5543] | 4606 [3428-5655] | 1.511 | 0.345 | 0.962 | 0.708 | 0.156 | 0.998 |
| Volumes are reported in $\text{mm}^3$. Significant differences are in bold. | | | | | | | | |

Total thalamus volume was not associated with motor composite scores in any groups (*p*>0.260) (Supplementary Table 4). In contrast, total lateral ventricle volume was associated with motor composite scores for control infants (*p*=0.040), but not in infants with CHD (*p*=0.829), or infants in the early preterm group (p=0.637) (Supplementary Table 4).

## 4. Discussion

Using a data-driven approach, we assessed intrathalamic morphometry in infants born preterm, infants with CHD, and typically developing controls. Preterm infants showed widespread differences across the thalamus compared to controls and infants with CHD. Similarly, GA at birth was associated with component weightings across the thalamus. In contrast, alterations in infants with CHD were confined to medial, ventricular-bordering, and some anterior and ventrolateral areas. Further, we show that weightings in the right posterior thalamus were associated with motor scores in early childhood in preterm infants born <32 weeks GA.

### 4.1 Intrathalamic morphometry in preterm infants

Preterm infants born <32 weeks GA showed alterations to structural covariance across the thalamus compared to controls, except the IC encompassing the left posterior thalamus. GA at birth was associated with weightings in the same components, suggesting disrupted development across the thalamus in preterm birth. In keeping with this finding, reduced thalamus volumes have been reported in preterm infants^8,11,43,44,59^, and have been associated with decreased cortical volume and altered microstructure of connecting white matter tracts^8^. Thalamocortical connectivity is also altered in preterm infants^15,16,94^. Together, these findings suggest preterm birth results in dysmaturation of thalamocortical systems. After adjusting for thalamus volume, IC weightings remained different in all components except the right posterior thalamus. This suggests that while differences in the right posterior thalamus may be driven by volumetric changes, preterm infants show alterations to thalamic morphometry beyond global size reductions across ventrolateral, medial, anterior, dorsal, and dorsal-posterior thalamic regions. Literature on volumetric reductions to specific regions of the thalamus in preterm infants is limited. Using voxel-based morphometry, Boardman and colleagues^58^ reported volumetric reductions in the mediodorsal thalamus in preterm infants with diffuse white matter injury. Procedural pain following preterm birth has also been associated with volumetric reductions to the lateral thalamus^41^. However, a recent study using diffusion MRI to parcellate the thalamus reported that relative volumes of intrathalamic clusters did not significantly differ between preterm and control infants^59^. Previous research has also shown alterations to thalamic shape in preterm infants^43,45,46^, and reduced surface area in ventrolateral, anterior, dorsomedial, and posterior thalamic regions^43^. It is possible that alterations to thalamic surface relate to the widespread morphometric differences we observe, however further research integrating combined analyses on thalamic shape, surface area, and intrathalamic morphometry in preterm infants is required. Nevertheless, our findings demonstrate widespread alterations in thalamic morphometric development in preterm infants.

### 4.2 Neurodevelopmental correlates of altered intrathalamic morphometry in preterm birth

We found that weightings in the IC4 were associated with motor abilities at 18 months in infants born <32 weeks GA. IC4, which encompasses the right posterior thalamus, includes part of the pulvinar nucleus. This nucleus is implicated in visual and attentional processing^95–97^ and evidence primarily from lesion studies suggests a broader role in visuomotor integration and in the selection and coordination of visually guided movements^98–101^. Although the pulvinar is most highly connected to primary visual and sensorimotor regions in infancy^13,102,103^, its function during infancy and early childhood is not fully understood. In a study of neonates born 37-43 weeks GA from the dHCP, morphometry in the right posterior thalamus was associated with a polygenic score for the age of onset of walking^104^. Taken together with our results, this suggests altered morphometry in this region may be a key antecedent of altered early motor development.

### 4.3 Intrathalamic morphometry in infants with CHD

Morphometric differences in infants with CHD were confined to medial, ventricle-bordering, and some ventrolateral and anterior regions of the thalamus. After adjusting for total thalamus volume, these differences remained, with additional alterations emerging in ventrolateral, medial, posterior, and ventricle-bordering tissues. Previous studies report reduced thalamus volume in this population^9,10,48,49^. Our findings suggest that morphometric differences to the thalamus are independent of global volumetric reductions. In keeping with our results, specific volumetric reductions to anterior and medial regions of the thalamus in infants with CHD have previously been reported^9^. Morphometric differences were also not explained by lateral ventricle volumes. Whilst lateral ventricle volumes are not enlarged in infants with CHD before surgery^9,34,48,105^, a previous study reported that the third ventricle is enlarged in infants with CHD before surgery^9^. All components that differed in infants with CHD included thalamic tissue bordering the third ventricle. It is therefore possible that the morphometric differences we observe may relate to enlargement of this structure. Beyond alterations in volumetry, one study has reported altered surface area and displacement of the thalamus in adolescence in CHD^106^. However, to our knowledge no studies have investigated surface area or displacement of the thalamus in the neonatal period in this population. It is possible that altered shape of CSF spaces bordering the thalamus accompany alterations in ventricle-bordering thalamic morphometry, although further research is required.

The mechanisms through which specific areas of the thalamus are altered in infants with CHD remains unknown. We found no association between IC weightings and CDO_2_ measured in the neonatal period. Previous studies in fetuses and infants with CHD report associations between reduced CDO_2_ and smaller brain and global thalamus volumes, as well as altered cortical morphometry^9,26,33,34^. However, other physiological factors in CHD may contribute to the thalamic alterations we report. For example, incidence and severity of placental pathology has also been associated with reduced deep gray matter volumes in fetal and infant CHD populations^32,107^. In addition, several genes are implicated in both cardiac and brain development^108–110^. Direct evidence linking placental pathology or genetic variation to thalamic morphometry in CHD remains limited and represents an important direction for future research.

Previous studies in this population have reported associations between reduced global thalamus volumes and cognitive ability at 22 months^26^ and IQ at 6 years^25^. However, in this study, no associations were observed between IC weightings and neurodevelopmental outcomes in infants with CHD. Intrathalamic differences in the CHD group observed here were primarily located in areas overlapping with medial and ventrolateral nuclei. Medial thalamic nuclei are implicated in higher-order executive functions including working memory, attentional control, and decision-making^111–114^, while ventrolateral nuclei play a central role in motor planning, coordination, and control^115–117^. Children with CHD are more likely to have impairments across these cognitive and motor functions at school-age^118–120^. However, these neurodevelopmental domains are not assessed in toddlerhood using the Bayley-III. Longitudinal studies assessing school age neurodevelopmental outcomes are required to determine whether early alterations to thalamic morphometry across medial and ventrolateral regions of the thalamus are associated with motor coordination and executive functions in children with CHD.

### 4.4 Comparisons between preterm infants and infants with CHD

Both early preterm and CHD populations showed altered thalamic morphometry compared to typically developing controls, indicating atypical thalamic development in both conditions. However, in direct comparisons, our results demonstrate distinct intrathalamic alterations in infants with CHD and preterm infants born <32 weeks GA. These findings align with emerging literature demonstrating unique brain alterations in these populations. Bonthrone and colleagues^51^ reported distinct changes in cortical scaling in CHD and preterm infants, while two recent studies have reported different patterns of atypical global connectivity between these populations^52,53^. Similarly, Wilson and colleagues^27^ showed differing patterns of altered structural covariance in a cohort of infants born preterm and with CHD overlapping with our study set, including in the whole thalamus. After adjusting for total thalamus volume, all but one component was no longer significantly different between **preterm** infants and infants with CHD. This suggests that volumetric differences in the thalamus may account for some differences in thalamic morphometry between groups. Infants born preterm can experience multiple adverse exposures including suboptimal intrauterine environment, inflammation, altered gut microbiota, respiratory distress, infection, pain, and stress^35–40,42^. Infants with CHD may experience an adverse intrauterine environment, however they are not exposed to the deleterious effects of early exposure to the extrauterine environment. It is possible that both conditions disrupt thalamic development, including reduced volumes, altered morphometry, and connectivity profiles, albeit due to different exposures and through differing biological pathways. Our findings suggest that early preterm birth could disrupt these thalamic developmental processes more severely, consistent with greater volume reductions and more widespread alterations to thalamic morphometry in comparison with controls.

### 4.5 Limitations

We acknowledge several limitations to our study. First, registration used a 40-week template, which may introduce subtle alignment biases for very preterm infants and infants with CHD with altered anatomy. Secondly, we did not assess differences in thalamic morphometry between the different types of CHD. Future work in large cohorts is needed to assess whether specific cardiac lesions are associated with different patterns of altered intrathalamic morphometry. Thalamic morphometry was also assessed prior to corrective cardiac surgery. As such, the impact of surgical intervention on intrathalamic morphometry, and its potential relation to neurodevelopmental outcomes is not known. Finally, neurodevelopmental outcomes captured prior to two years may not be able to capture certain domains of altered neurodevelopmental outcomes in these population, such as higher-order motor abilities and some executive functions. Future studies should consider neurodevelopmental outcomes at school-age to address how thalamic alterations in infancy relate to these functional domains.

## 5. Conclusion

By applying a data-driven parcellation approach we show that intrathalamic morphometry is altered in both preterm infants and neonates with CHD. Preterm infants showed widespread alterations in the thalamus, and differences in the right posterior thalamus were associated with motor ability in early life. Alterations in infants with CHD were confined to medial, ventricular-bordering and some ventrolateral and anterior regions and were not associated with early neurodevelopmental outcomes. Our results suggest non-identical alteration to thalamic morphometry in infants born preterm and with CHD, and that preterm birth may alter thalamic morphometry to a greater extent than CHD.

## 6. Author Contributions

B.C., A.F.B., and S.J.C., conceived and designed the study, B.C. processed, analyzed and interpreted the data and drafted the manuscript. A.F.B., B.G.E., and M.E.M.v.d.M. assisted with data analysis. D.C., C.K., and A.T.M.C. collected the data, M.E.M.v.d.M., D.C., and A.T.M.C. curated the data. M.A.R. reviewed all images. K.P. and J.S. provided clinical assistance and cardiology input. S.W., J.O.M., and C.N. assisted in development of the methodology. J.V.H provided technical assistance for MR image acquisition. A.D.E., J.V.H, A.F.B, and S.J.C acquired funding. A.F.B. supervised processing, analysis, interpretation and manuscript editing. All authors reviewed and edited the manuscript.

## 7 Acknowledgments

This research was funded by the British Heart Foundation [PG/24/11831; FS/15/55/31649], Medical Research Council (MRC) UK [MR/L011530/1; MR/V002465/1], and Action Medical Research [GN2630]. The Developing Human Connectome Project was funded by the European Research Council under the European Union’s Seventh Framework Program [FP7/20072013]/European Research Council Grant agreement no. 319456. This research was supported by the Wellcome Engineering and Physical Sciences Research Council Centre for Medical Engineering at King’s College London [WT 203148/Z/16/Z], and by the NIH Research (NIHR) Biomedical Research Centre based at Guy’s and St Thomas’ National Health Service (NHS) Foundation Trust and King’s College London. J.O. is supported by a Sir Henry Dale Fellowship jointly funded by the Wellcome Trust and the Royal Society [206675/Z/17/Z]. A.D.E., J.O.M, and M.A.R. receive funding from the MRC Centre for Neurodevelopmental Disorders, King’s College London [MR/N026063/1]. The views expressed are those of the authors and not necessarily those of the NHS, the NIHR, or the Department of Health. We would like to thank the families who participated in this study. We also thank our research radiologists, our research radiographers, and our neonatal scanning team. In addition, we thank the staff from the St Thomas’ Hospital Neonatal Intensive Care Unit; the Evelina London Children’s Hospital Fetal and Paediatric Cardiology Departments; the Evelina London Paediatric Intensive Care Unit; and the Centre for the Developing Brain at King’s College London.

## 9. Data Availability Statement

The data from the dHCP 3^rd^ neonatal release is publicly available (https://nda.nih.gov/edit_collection.html?id=3955). Anonymized derived data from 50 infants with CHD acquired under ethics 21/WA/0075 and infants from dHCP (n = 485), as well as analysis scripts, are available from the corresponding author. We do not have ethical permission for sharing data for 57 infants with CHD acquired under ethics 07/H0707/105.

