## Supplementary materials for "Intrathalamic morphometry in infants with congenital heart disease and infants born preterm"

**Supplementary Table 1. Total variance explained for different numbers of independent components (ICs) selected**

| **Number of ICs** | **Total Variance Explained** |
| --- | --- |
| 2 | 0.1148046 |
| 3 | 0.1385757 |
| 4 | 0.1307201 |
| 5 | 0.1484610 |
| 6 | 0.1312736 |
| 7 | 0.1548877 |
| 8 | 0.1448772 |
| 9 | 0.1539808 |
| 10 | 0.1500468 |

**Supplementary Table 2. Association between cerebral delivery of oxygen (CDO_2_) and IC weights**

|  | **b** | ***p*** | ***p*_FWE_** |
| --- | --- | --- | --- |
| **IC1** | 0.000038 | 0.957 | 1.000 |
| **IC2** | -0.000337 | 0.431 | 1.000 |
| **IC3** | -0.000830 | 0.221 | 1.000 |
| **IC4** | -0.000080 | 0.883 | 1.000 |
| **IC5** | 0.000141 | 0.734 | 1.000 |
| **IC6** | -0.000928 | 0.073 | 0.514 |
| **IC7** | 0.000457 | 0.439 | 1.000 |
| **IC8** | 0.000708 | 0.056 | 0.444 |

*adjusting for PMA at scan, PNA at scan, birthweight, sex and whether the infant was part of a multiple birth.*

**Supplementary Table 3. Association of weightings in IC4 with motor composite scores**

|  | **Group** | **b** | **p** | **permuted p** |
| --- | --- | --- | --- | --- |
| (i) Motor score ~ IC4 \| Covariates | Control | -0.102 | 0.633 | 0.634 |
|  | CHD | -0.289 | 0.588 | 0.581 |
|  | Early Preterm | -1.911 | **0.037** | **0.037** |
| (ii) Motor score ~ IC4 + Thalamus volume \| Covariates | Control | -0.101 | 0.635 | 0.639 |
|  | CHD | -0.378 | 0.496 | 0.489 |
|  | Early Preterm | -2.070 | **0.024** | **0.024** |
| (iii) Motor score ~ IC4 + Lateral Ventricle volume \| Covariates | Control | -0.212 | 0.329 | 0.331 |
|  | CHD | -0.277 | 0.615 | 0.608 |
|  | Early Preterm | -1.894 | **0.044** | **0.043** |

*adjusting for PMA at scan, PNA at scan, birthweight, sex, IMD and whether the infant was part of a multiple birth.*

**Supplementary Table 4. Association of thalamus or lateral ventricle volume with motor composite scores**

|  | Group | **b** | **p** | **permuted p** |
| --- | --- | --- | --- | --- |
| (i) Motor score ~ Thalamus volume \| Covariates | Control | 0.000714 | 0.376 | 0.378 |
|  | CHD | 0.000797 | 0.671 | 0.667 |
|  | Early Preterm | 0.00148 | 0.270 | 0.270 |
| (ii) Motor score ~ Lateral Ventricle volume \| Covariates | Control | -0.000754 | **0.035** | **0.040** |
|  | CHD | 0.000165 | 0.835 | 0.829 |
|  | Early Preterm | 0.000330 | 0.655 | 0.637 |

*adjusting for PMA at scan, PNA at scan, birthweight, sex, IMD and whether the infant was part of a multiple birth. Significant associations are in bold.*

**Supplementary Table 4. Comparison of thalamus and lateral ventricle volume between infant groups**

|  | CHD vs Control | | | Early Preterm Vs Control | | | CHD vs Early Preterm | | |
| --- | --- | --- | --- | --- | --- | --- | --- | --- | --- |
|  | t | p | perm p | t | p | perm p | t | p | perm p |
| Thalamus Volume | 5.972 | **<0.001** | **<0.001** | 33.548 | **<0.001** | **<0.001** | 13.828 | **<0.001** | **<0.001** |
| Lateral Ventricle Volume | 1.511 | 0.131 | 0.131 | 0.962 | 0.336 | 0.336 | 0.156 | 0.876 | 0.875 |

*adjusting for PMA at scan, birthweight, sex, IMD and whether the infant was part of a multiple birth. Comparisons between control and infants with CHD were additionally adjusted for PNA at scan. Significant differences are in bold.*

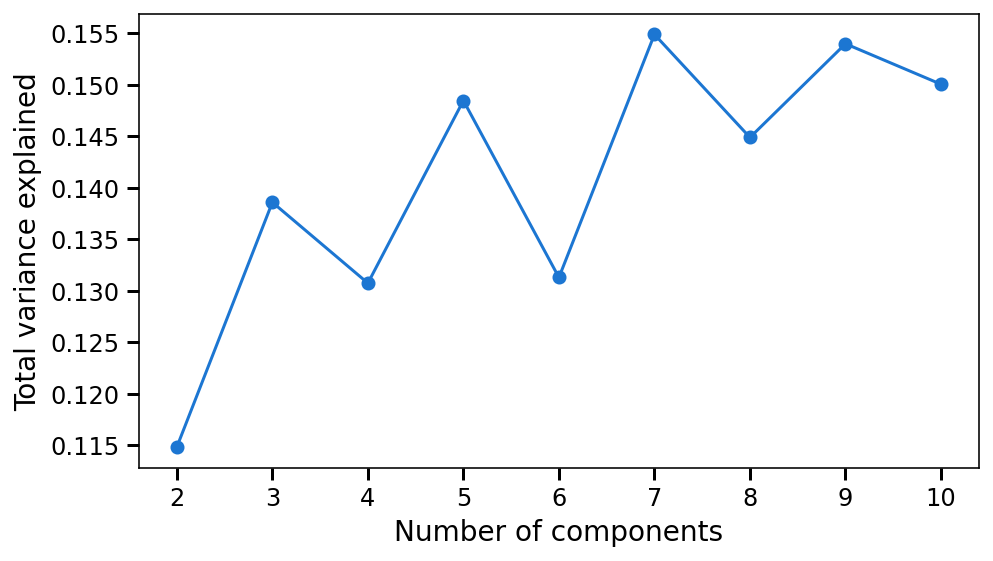

**Supplementary Figure 1.** Total variance explained for different numbers of independent components (ICs).
